# Faithful Diagnoses: Association of Socioeconomic Status with Diagnoses of ASD and ADHD in Girls

**DOI:** 10.1101/2025.05.11.25327397

**Authors:** Armaan Mehta

## Abstract

**Purpose:** Autism (ASD) and attention-deficit/hyperactivity disorder (ADHD) are chronic developmental disorders that have drastically increased in prevalence and co-occurrence over the past two decades. In previous research, ADHD has been correlated with low income and autism with high income, but it is uncertain how gender affects this. This analysis tests each of these income correlations, socioeconomic status (SES) confounders, and income modality against both diagnoses in girls.

**Methods:** Cross-sectional data from the 2020 National Survey of Children’s Health was analyzed. Multiple logistic regressions tested for correlations between each diagnosis and income, bimodal diagnosis patterns were tested at low and high income, and posttest analyses compared the respective odds ratios against both diagnoses.

**Results:** This analysis was unable to detect any of the previously researched relationships between diagnosis in girls and income, nor a significant bimodal correlation of either diagnosis with income. Low income was linearly associated with autism diagnosis (odds ratio: 0.997, *p* = 0.011) while ADHD was not correlated with income. A peak in both diagnoses was observed near 138% of the federal poverty limit (FPL). Compared with White girls, only Hispanic Asian girls showed higher odds of autism in this analysis (OR: 8.2, *p =* 0.023).

**Conclusion:** Previously observed trends between SES and ASD or ADHD did not appear specifically in girls. Instead, girls experience income, SES, and ethnoracial trends unique to them. A peak in diagnoses near 138% FPL suggests that expanded Medicaid eligibility may play a larger role for autism diagnosis in girls than expected.

## Introduction

Attention deficit hyperactivity disorder (ADHD) and autism spectrum disorder (ASD) are chronic disorders commonly diagnosed in children. ADHD prevalence has doubled from under 6% to over 10% over the past two decades while autism has tripled from 0.67% to 2.7% (Akinbami et al., 2011; Zablotsky & Black, 2020; Maenner et al., 2023). Both are diagnosed in males much more than females (Mahendiran et al., 2019a), though, how much of this disparity is due to neurobiological differences versus social gender norms is still widely debated (Lai & Szatmari, 2020; Mowlem et al., 2019). Researchers have proposed that girls are underdiagnosed due to ignored or missed symptomology due to their social gender behavior patterns; for example, the “Female Autism Phenotype” (Hull et al., 2020). Even within social behavior comparisons, studies have found inconsistent evidence comparing girls and boys (Mahendiran et al., 2019a; Mahendiran et al., 2019b). These comparisons frequently had limited data on girls and did not include ethnoracial or socioeconomic comparisons.

Lower socioeconomic status (SES) increases exposure to environmental factors that contribute to mental disorders, both chronic and acute, and especially in children (Peverill et al., 2020). Low SES has a well-established correlation with ADHD (Spencer et al., 2022, Russell et al., 2016) while the opposite is true for autism (Dickerson et al., 2017). For ADHD, this correlation is slightly reduced by parental history of the diagnosis, likely due to epigenetics and awareness (Rowland et al., 2018). Previous research has also shown that access to diagnosticians through Medicaid coverage can be a main driver of increased ADHD diagnoses in low-income children, (Chorniy et al., 2018). Some researchers have suggested that a similar access bias may be present in autism diagnoses as well. Specifically, wealthier counties and states may have better access to diagnosticians allowing for more diagnoses (Dickerson et al., 2017; Thomas et al., 2012). It is unclear and understudied how sub-demographics drive correlations with SES, or if these trends are affected by gender.

While these socioeconomic trends with autism and ADHD continue to be developed, additional research is needed explore these trends in girls. This paper evaluates the relationship between SES and both diagnoses in girls and the differences in SES between the two diagnoses in girls. This paper also investigates the demographic trends of girls diagnosed with autism and ADHD. Lastly, this paper tests for modality in the relationship between SES and both diagnoses in girls, with potential peaks in diagnoses at high and low incomes.

## Methods

### Data Source and Study Sample

This study is a secondary analysis of the publicly available 2020 National Survey of Children’s Health (NSCH), a nationally representative cross-sectional survey on the health and well-being of 42,777 children ages 0-17 in the United States. This analysis is restricted to 20,673 families with girls aged 0-17, representing 35,582,224 girls across the U.S. total.

The survey is designed to assess children’s health, healthcare, social and family environment, livelihood, and behaviors. Each participant was given one of three surveys based on the sampled child’s age, according to the ranges of 0-5, 6-11, and 12-17 years old. The healthcare section included questions about medication, therapeutics, diagnoses, and included questions on ADHD and autism diagnoses.

The 2020 NSCH does not contain variables for gender or related identities. This analysis used the “sex” variable to indicate sex and gender identity.

### Study Measures

#### Dependent Variables

*Sex and gender:* If a respondent answered “female” to the question “What is this child’s sex?” they were categorized as being a girl, or female.

*ADHD diagnosis:* If a respondent answered “yes” to the question, “Has a doctor or other health care provider EVER told you that this child has Attention Deficit Disorder or Attention-Deficit/Hyperactivity Disorder, that is, ADD or ADHD?” they were categorized as having an ADHD diagnosis.

*ASD diagnosis:* If a respondent answered “yes” to the question, “Has a doctor or other health care provider EVER told you that this child has Autism or Autism Spectrum Disorder (ASD)?” they were categorized as having an autism diagnosis.

#### Key Independent Variable

Data on annual income were collected by asking the interviewee, “Think about your total combined family income in the last calendar year… what is that amount before taxes?” Responses were stored as a percentage of the federal poverty level (FPL), ranging from 50% to 400%.

#### Covariates/Confounders

The dataset includes a variable on race with the following six categories: White, Black, American Indian/Alaskan Native (AIAN or “Native American”), Asian, Native Hawaiian/Pacific Islander (NHPI), and two or more races. A variable on ethnicity indicates whether the child is of Hispanic, Latino, or Spanish origin. These two variables were combined to form a single variable of Hispanic (H) for each race, and non-Hispanic (NH) for each race. Interviewees were asked “How old is this child?” and given the ability to specify the child’s age in years or months. There are 3 cases of ADHD in children under 2 years old, denoted with months. These three cases of ADHD were included as ages 0 and 1 years in this analysis. The final age variable is a discrete (integer) variable that ranges from 0 to 17.

To obtain an estimate of the independent association of family income and the two dependent variables, a variable on health insurance coverage was included with the categories “Public only,” “Private only,” “Private and public,” and “Not insured.” Private insurance included government-related plans, such as TRICARE for the military, while all social insurance programs (e.g., Medicaid) were captured under public only.

Interviewees were asked “How many children are in [the] household?” and given response options of 1, 2, 3, or 4+ children. The final variable is an ordinal, discrete (integer) variable that ranges from 1 to 4 or more.

### Statistical Analysis

Two bivariate χ^2^ tests were performed on each explanatory variable. Once to test correlation of the variable with autism, and then to test correlation with ADHD. Table 1 describes the proportion of girls by the variable categories, and then by diagnosis within these categories. P values are shown for the Pearson χ^2^ coefficient, indicating if the differences between the proportion of girls with autism and ADHD within a demographic category are significant.

**Table 1.**
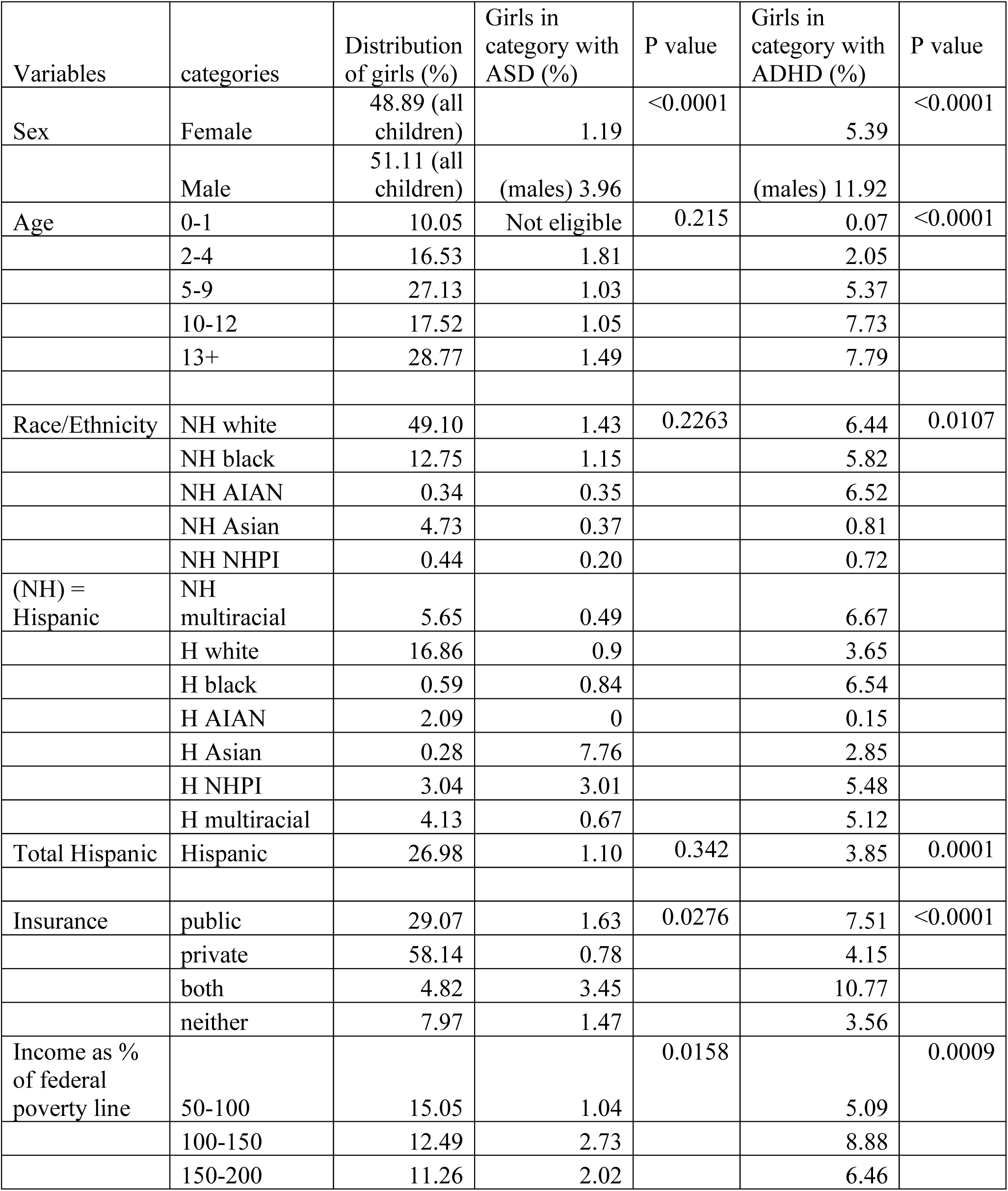
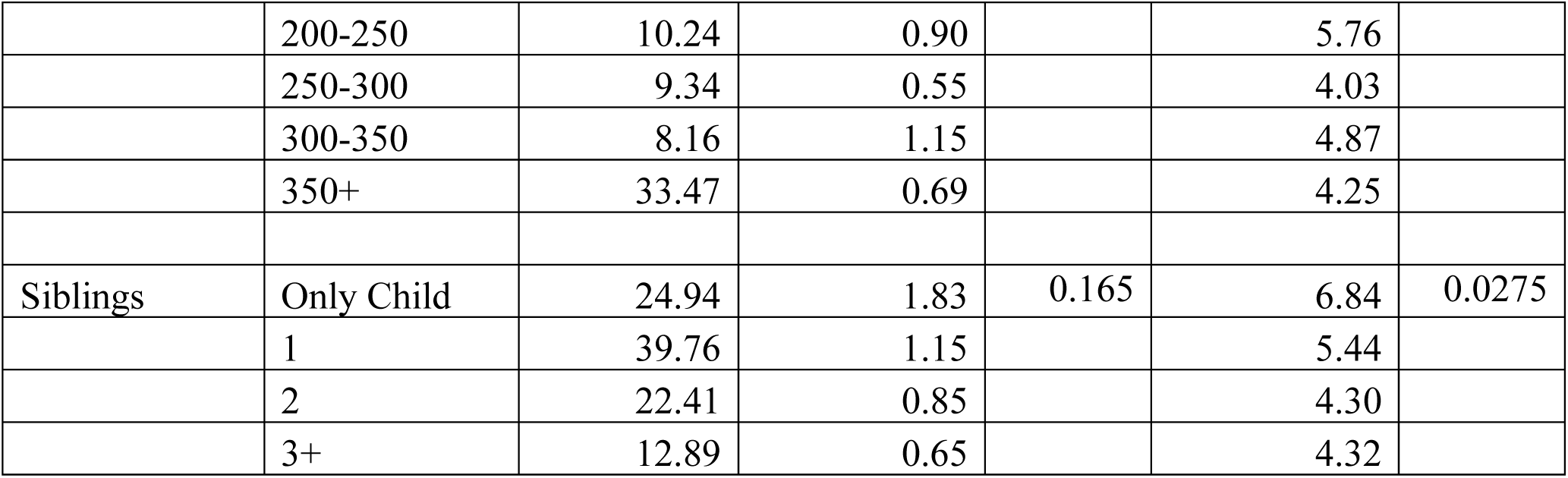

Two groups of logistic regressions were performed, one predicting autism and one predicting ADHD. For each, an unconfounded linear regression was performed (Table 2), a linear regression with confounders was performed (Table 3), and a quadratic regression with confounders was performed (Table 4). Income (FPL) was used as the main explanatory variable and income squared as the quadratic explanatory variable. Age, race, ethnicity, insurance type, and number of siblings were included as confounding explanatory variables. The results are shown as odds ratios (OR) for each variable, with associated *p* values for the regression.

**Table 2.**

| Measure | Range | Univariate<br>ASD OR | p | Univariate<br>ADHD<br>OR | p |
| --- | --- | --- | --- | --- | --- |
| F statistic | | F(1, 42590) = 8.72<br>$p = 0.0031$ | | F(1, 42574) = 10.05<br>$p = 0.0015$ | |
| income | 50-400 | 0.997 | 0.0031 | 0.998 | 0.0015 |

**Table 3.**

| Measure | Range | Model 1:<br>ASD OR | p | Model 2:<br>ADHD<br>OR | p |
| --- | --- | --- | --- | --- | --- |
| F Statistic | | F(16, 42186) = 7.93<br>$p < 0.0001$ | | F(17, 42248) = 18.99<br>$p < 0.0001$ | |
| income | 50-400 | .9971 | 0.011 | .999 | 0.207 |
| Income squared |  | NA | NA | NA | NA |
| age | 0-17 | 1.023 | 0.565 | 1.125 | <0.001 |
| race | NH white | 1 |  | 1 |  |
|  | NH black | .505 | 0.180 | .585 | 0.013 |
|  | NH AIAN | .155 | 0.039 | .725 | 0.508 |
|  | NH Asian | .233 | 0.037 | .103 | <0.001 |
|  | NH NHPI | .117 | 0.053 | .108 | 0.008 |
|  | NH multiracial | .281 | 0.009 | .952 | 0.829 |
|  | H white | .431 | 0.111 | .390 | <0.001 |
|  | H black | .328 | 0.199 | .539 | 0.154 |
|  | H AIAN | NA | NA | .011 | <0.001 |
|  | H Asian | 8.187 | 0.023 | .617 | 0.520 |
|  | H NHPI | 1.151 | 0.901 | .633 | 0.538 |
|  | H multiracial | 0.338 | 0.127 | .686 | 0.427 |
| insurance | public | 1 |  | 1 |  |
|  | private | .598 | 0.282 | .453 | <0.001 |
|  | both | 2.539 | 0.173 | 1.425 | 0.246 |
|  | neither | 0.961 | 0.941 | .395 | 0.001 |
| siblings | 1-4 | .618 | 0.019 | .825 | 0.011 |

**Table 4.**

| Measure | Range | Model 3:<br>ASD OR | p | Model 4:<br>ADHD<br>OR | p |
| --- | --- | --- | --- | --- | --- |
| F Statistic | | F(17, 42185) = 8.41<br>$p < 0.0001$ | | F(18, 42247) = 18.09<br>$p < 0.0001$ | |
| income | 50-400 | 1.005 | 0.347 | 1.000 | 0.978 |
| Income squared |  | .999984 | 0.133 | .999998 | 0.673 |
| age | 0-17 | 1.022 | 0.578 | 1.125 | <0.001 |
| race | NH white | 1 |  | 1 |  |
|  | NH black | .523 | 0.200 | .587 | 0.013 |
|  | NH AIAN | .155 | 0.038 | .724 | 0.509 |
|  | NH Asian | .236 | 0.038 | .103 | <0.001 |
|  | NH NHPI | 0.114 | 0.052 | .108 | 0.008 |
|  | NH multiracial | .275 | 0.009 | .950 | 0.823 |
|  | H white | .439 | 0.116 | .391 | <0.001 |
|  | H black | .340 | 0.214 | .543 | 0.158 |
|  | H AIAN | NA | NA | .011 | <0.001 |
|  | H Asian | 8.196 | 0.023 | .618 | 0.520 |
|  | H NHPI | 1.281 | 0.826 | .639 | 0.548 |
|  | H multiracial | .321 | 0.113 | .682 | 0.427 |
| insurance | public | 1 |  | 1 |  |
|  | private | .618 | 0.286 | .454 | <0.001 |
|  | both | 2.444 | 0.180 | 1.417 | 0.249 |
|  | neither | .960 | 0.939 | .395 | 0.001 |
| siblings | 1-4 | .617 | 0.020 | .826 | 0.011 |

In addition, an axis of symmetry test was performed on the regression coefficients for income and income squared. The axis of symmetry tests the size and location of a quadratic, or U-shaped curve between income and the diagnoses, if it exists (Table 5). This was calculated using the -b/2*a^2^ formula, or specific to this analysis: -income/(2*income^2^).

**Table 5.**

| Diagnosis | Value of symmetry | p | 95% CI lower | 95% CI upper | Direction |
| --- | --- | --- | --- | --- | --- |
| ASD (girls) | 142.21 | 0.027 | 16.05 | 268.38 | Peak |
| ADHD (girls) | 14.77 | 0.976 | -962.46 | 991.99 | Peak |
| ASD (boys) | 233.24 | 0.072 | -20.65 | 487.12 | Peak |
| ADHD (boys) | 266.42 | <0.001 | 212.61 | 320.23 | Trough |

Using the “suest” command available in the Stata software, Wald tests and directional Wald tests were performed on income and income squared to test if the odds ratios of autism were greater or less than the odds ratios for ADHD when income was changed (Table 6). This method of postestimation using Wald tests follows STATA’s recommended protocol for hypothesis testing in postestimation (<u>StataCorp, 2021</u>).

**Table 6.**

| Variable for ASD | Comparison | Variable for ADHD | F test | P value |
| --- | --- | --- | --- | --- |
| Income model 1 | Equal to | Income model 2 | 2.98 | 0.084 |
| Income model 1 | Greater than | Income model 2 |  | 0.042 |
| Income model 3 | Equal to | Income model 4 | 0.94 | 0.333 |
| Income model 3 | Greater than | Income model 4 |  | 0.833 |
| Income squared | Equal to | Income squared | 1.89 | 0.170 |
| Income squared | Greater than | Income squared |  | 0.085 |

All statistical analyses were conducted with the Stata/SE 18 software program. The “svy” prefix and “svyset” command was used to account for the stratification, weighting, and special needs sampling design according to US Census Guide (U.S. Census, 2022). All estimations used a *p* value of less than .05, or 95% confidence intervals, for statistical significance. All code is available in the Supplementary STATA File and at https://github.com/amehta63/NSCH-ASD-ADHD-analysis. All data is available at https://www.census.gov/programs-surveys/nsch/data/datasets.html.

## Results

Table 1 shows the distribution of each variable within the subpopulation of girls aged 0 to 17 in the U.S., as well as the percentage of each explanatory variable category that are both female and diagnosed with autism or ADHD. Weighted, the NSCH analytic sample is representative of 35.6 million girls. Ten percent of these girls are under the age of 2, and ineligible to be diagnosed with autism, but had a 0.07% prevalence of ADHD. While age is significantly correlated with ADHD diagnoses (*p <* 0.001), it is not correlated with autism diagnoses (*p* = 0.2154).

Table 1 also shows that race and ethnicity do not differ significantly among autism diagnoses (*p* = 0.2263 and *p* = 0.342, respectively), however, they do with ADHD diagnoses (*p* = 0.0107 and *p* = 0.0001, respectively). NH Asians and NH NHPI had the lowest rate of ADHD among non-Hispanic girls, at 0.81% and 0.72% respectively (*p* = 0.0001). Among Hispanics, American Indian and Alaskan Native girls had the lowest rate of ADHD at 0.15%. NH Black girls had slightly lower rates of ADHD than NH White girls (5.82% vs 6.44%, *p =* 0.0107), while Hispanic Black girls had almost double the rate of ADHD as Hispanic White girls (6.54% vs 3.65%, *p =* 0.0107).

Girls with a combination of public and private insurance have a higher prevalence of both autism and ADHD compared to girls with either or no insurance. Specifically, girls with public, private, or no insurance had an ADHD prevalence of 7.51 %, 4.15%, and 3.56% respectively, while girls with combination insurance had a rate of 10.77% (*p* < 0.0001). Girls with public, private, or no insurance had an autism prevalence of 1.63%, 0.78%, and 1.47%, respectively, while girls with combination insurance had a rate of 3.45% (*p =* 0.0276). Girls with private insurance had an especially low rate of autism at 0.78% (*p =* 0.0276).

Sibling count was significantly correlated with ADHD (*p* = 0.0275) but not autism *(p* = 0.165). In both diagnoses girls who were an only child had the highest rate of diagnosis; for ADHD specifically, this was 6.84%.

Differences in income were significantly correlated with both autism and ADHD diagnoses. Girls in households with incomes between 100% and 150% of the federal poverty line had the highest prevalence of autism and the second highest prevalence of ADHD at 2.73 % and 8.88%. A line histogram of diagnosis prevalence vs income level using 30% FPL bins in both girls and boys can be seen in *Figure 1.* The histograms for ADHD and autism in girls both have a mode between 135% and 140% FPL, with smaller bins centered at 138%.

**Figure 1:**
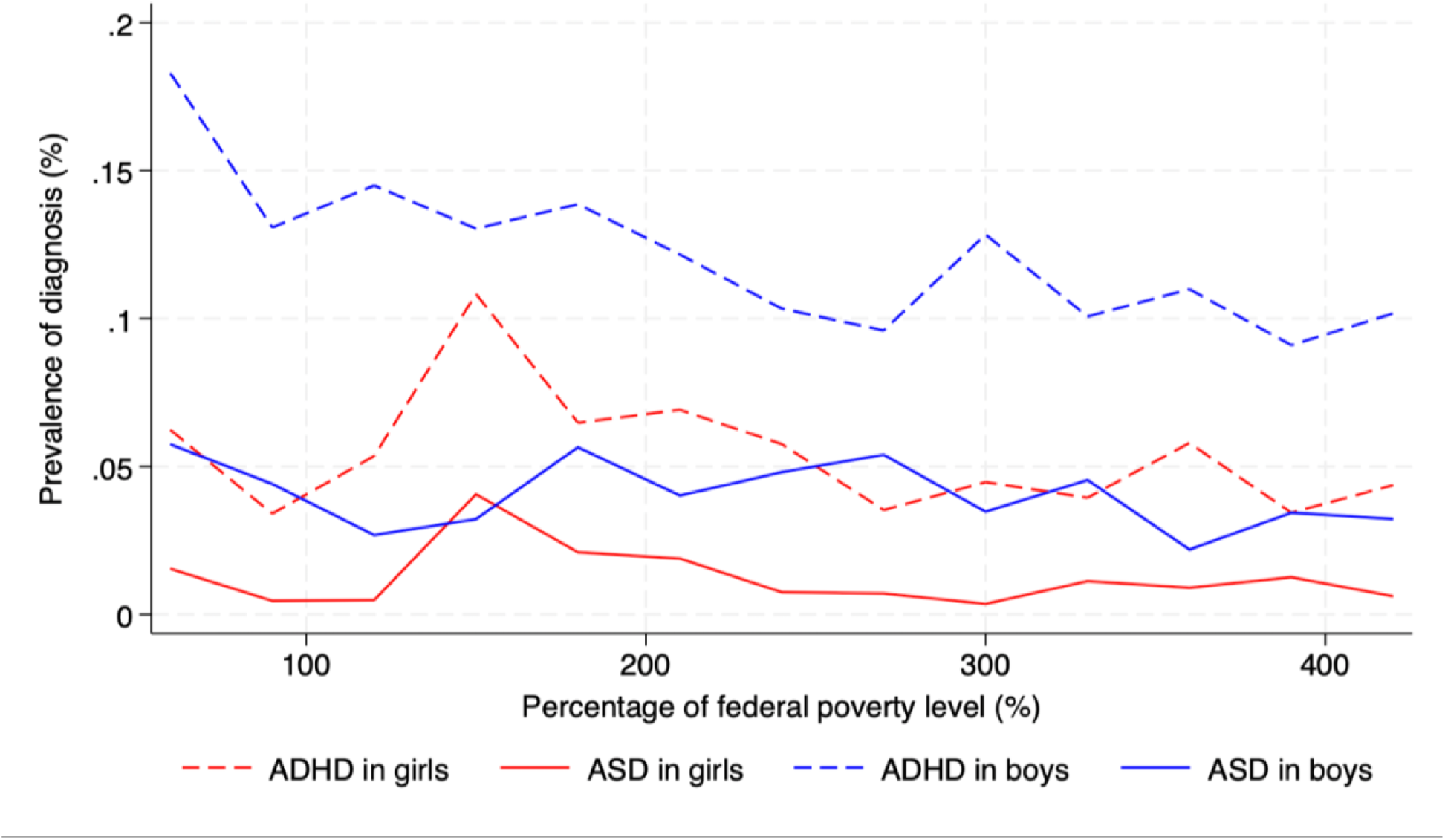
Prevalence of ASD and ADHD by Income Level

Table 2 shows a univariate logistic regression of income predicting autism and ADHD diagnoses. Income alone predicted autism diagnoses with an OR of 0.997 (*p =* 0.003), and ADHD diagnoses with an OR of 0.998 (*p =* 0.002).

### Multivariate Results

Tables 3 and 4 show the ORs for multivariate logistic regression models predicting autism and ADHD. Models 1 and 3 predict autism with model 1 using only a linear term for income and model 3 using both a linear and quadratic term for income. Models 2 and 4 predict ADHD with model 2 using only a linear term for income and model 4 using both a linear and quadratic term for income. Although all four regressions were significant, only one of the regressions (model 1) showed income level as a significant predictor of diagnosis, though it was small in magnitude. Specifically, income linearly predicted autism diagnoses with an OR of 0.997 (*p* = 0.011). A Wald test of equivalence was performed to test if autism coefficients were statistically unlikely to equal ADHD coefficients (Table 6). Only the income coefficient in model 1 for autism was shown to be significantly greater than the equivalent coefficient for ADHD (*p* = 0.042).

### Bimodal “U-shape” Tests

To test for peaks in diagnoses due to income levels, the quadratic models were tested for a change in direction, or a parabolic “U shape” in the curve (Table 5). Peaks in the curve were considered axes of symmetry and were evaluated for what income level they occurred at. The axis of symmetry for income as a predictor of autism was centered at a level of 142.21 (*p* = 0.027, 95% CI: 16.05 ∼ 268.38). Since the center falls within the range of income values (50 – 400) this model predicted a peak in the relationship between income and autism at 142% of the federal poverty level.

The axis of symmetry for the predictor of ADHD was centered at an income level of 14.77 (*p* = 0.976, 95% CI: -962.46 ∼ 991.99). This test failed to reject the null hypothesis that ADHD does not have a modal relationship with income level.

### Confounders

Autism was significantly predicted by several races and the number of siblings a girl had. Hispanic Asian girls were the only race with higher odds of being diagnosed with ADHD than NH White girls (OR = 8.2, *p =* 0.023). In contrast, NH Asian girls had a much lower odds of being diagnosed with ADHD (OR = 0.23, *p =* 0.037). The odds of an autism diagnosis were lower in American Indian/Alaskan Native girls (OR = 0.15, *p* = 0.039), Native Hawaiian/Pacific Island girls (OR = 0.12, *p =* 0.053), and multiracial girls (OR = 0.28, *p =* 0.009) when compared to NH White girls. For each sibling the odds of being diagnosed with autism were 38% lower (OR = 0.62, *p* = 0.019).

In model 2, all non-Hispanic, non-White races were less likely to be diagnosed with ADHD than NH White girls. NH Black girls (OR = 0.58, *p =* 0.013), NH Asian girls (OR = 0.10, *p <* 0.001), NH Native Hawaiian/Pacific Island girls (OR = 0.11, *p =* 0.008), and Hispanic White girls (OR = 0.39, *p <* 0.001) all had significantly lower likelihood of being diagnosed with ADHD. Hispanic Asian girls had the lowest odds of diagnosis, with an OR of 0.011 (*p <* 0.001). For each additional year of age, the odds of being diagnosed with ADHD are 53% higher (OR = 1.125, *p <* 0.001). This is likely an artifact of the diagnostic criteria of ADHD, which has thresholds at 7 years old (DSM IV) and 12 years old (DSM-5). When children under 12 years old are excluded, the statical effect of age disappears, but all ages are included in this analysis to maintain the integrity of the income model. Private insurance and no insurance significantly lowered the odds of being diagnosed with ADHD (OR of 0.45 and 0.40, *p* ≤ 0.001, respectively) compared to public insurance. Like autism, siblings decreased the odds of being diagnosed with ADHD with each sibling lowering the likelihood by 45% (OR = 0.83, *p* = 0.011).

The remaining confounding variables were not associated with autism or ADHD diagnoses.

## Discussion

This study documents current trends in socioeconomic status, income levels, race, ethnicity, and age for autism and ADHD diagnosed girls. As current autism and ADHD research remains predominantly focused on boys, this study adds to a large gap in what is known about the demographics of these diagnoses in girls.

Previous research that focused on children of all genders established a correlation between low SES and ADHD and a correlation between high SES and autism (Dickerson et al., 2017; Spencer et al., 2022; Rong et al., 2021). This paper hypothesized that both trends would be present in a sub-analysis of girls. This would have created a bimodal relationship between SES and diagnosis, where both ADHD and autism would have increased diagnoses at high and low SES, and decreased diagnoses at middle SES. This analysis did not find either the previously researched correlations, or the hypothesized ones, among girls in the United States. This analysis found significant differences in diagnoses between income groups, but no overall trend between income and diagnosis of ADHD in girls was found. Only a slight negative correlation between low SES and autism in girls was found (OR of 0.997, *p* = 0.018). This odds ratio of near 1 suggests that this correlation was likely only due to a peak in diagnoses at incomes close to 140% of the federal poverty level.

Although multiple confounding variables were shown to have significance in this model, they did not further elucidate the SES relationship expected. A comparison of the univariate and multivariate models shows that, although race and ethnicity are significant predictors of diagnosis, they did not significantly alter the effect of income. Previous research on children of all genders had found that Black children and Hispanic children have increased in diagnosis rates and had similar or elevated rates of autism diagnoses compared to White children in 2021 (Pham et al., 2022). Neither of these correlations were found among girls. Further, although these two groups are associated with lower socioeconomic status, they did not alter the SES model in this analysis. Previous analyses of ADHD in children of all genders showed slightly lower rates of ADHD in Hispanic children and slightly higher rates of ADHD in Black children when compared to White children (Zablotsky & Alford, 2020). These previous analyses also showed a dramatic increase in ADHD rates for children living under 100% of the federal poverty limit.

This analysis did not find either this racial trend or the income trend in ADHD in girls. Rather, both Hispanic girls as a whole and NH Black girls had lower rates of ADHD than NH White girls, NH Native American girls, and multiracial girls. This analysis also showed that the OR of autism in Hispanic Asian girls compared to NH White girls was especially high (OR = 8.2, *p =* 0.023). The choice to maintain the breakdown of race and ethnicity of Hispanic girls was particularly important in measuring this.

This analysis found a dramatic increase in both autism and ADHD in girls between 135% and 140% of the federal poverty limit. Two different parts of this analysis detected this peak. The line histogram in *Figure 1* shows a peak in both diagnoses in girls centered on 138% of the federal poverty limit. Similarly, the higher-order logistic model for autism in girls found a change in direction, or “inverted U-shape” in income at 142% of the federal poverty level.

Neither peak was present in boys.

This peak in correlation between income and diagnosis suggests that expanded Medicaid eligibility plays a significant role in the diagnosis of ADHD and autism in girls. Previous research has found that Medicaid enrollment is associated with both autism and ADHD diagnoses (Engelhard et al., 2020). Similarly, expanded access to diagnosticians through Medicaid drove significant amounts of the increase in ADHD diagnoses from 2010 to 2013 (Chorniy et al., 2018). It is likely that Medicaid eligibility and access to diagnosticians plays a similar role for girls and their families. Since this peak was not seen in boys, research is needed to further identify the effect of Medicaid eligibility on sex and gender differences.

Accurate demographic knowledge of autism and ADHD are crucial to future work with these diagnoses. This study has introduced large departures from previously measured demographics trends when specifically considering girls. This is especially true of racial, ethnic, and income trends. This study lays the foundation to expand future research on the demographic-specific needs of girls, as well as considerations for understanding income-related trends in diagnoses, such as how girls’ access to autism and ADHD diagnoses are affected by Medicaid eligibility.

### Strengths and Limitations

The greatest strength of this study is the generalizability of the analysis, and the granularity of its demographics. The NSCH is a highly generalizable dataset, as it is designed to be a representative sample of the entire U.S. It is also one of a few datasets that include diagnoses for both autism and ADHD, as well as socioeconomic status indicators for such a wide population. This combination allows for the highly generalizable demographic analysis in this paper.

The largest limitation of this study is the NSCH’s caps on income at 50% and 400% of the federal poverty level. Due to this, income analysis cannot be expanded outside this range. Unfortunately, there are likely important trends below 50% FPL and above 400% FPL that contribute to autism and ADHD that cannot be studied here.

The NSCH also does not include data on the gender identity of the reported child. This miscategorizes an important population of genderqueer children who may experience SES and mental health diagnoses in different ways than cisgender children. It is likely that this analysis only included assigned female at birth children, which may have skewed the data.

Although this analysis focuses on confounders of diagnosis of autism and ADHD in girls, it did not include a comparison of these confounders between girls and boys. This study also did not consider diagnostic desirability. Some research has shown that, historically, external perceptions of girls with ADHD are far worse than their male counterparts (Eisenberg & Schneider, 2007). Some authors have theorized that new social media trends have reduced the stigmatization of ADHD and autism (McDermott, 2022). Future analyses that include measures of diagnostic desirability will help elucidate these trends.

Lastly, this study was not able to successfully predict co-occurrence or differential diagnosis of autism and ADHD. Until the fifth edition of the Diagnostic and Statistical Manual of Mental Disorders (DSM-5, published in 2013), co-occurrence of ADHD and autism (formerly “Pervasive Developmental Disorders”) was not recognized (APA, 2000; APA, 2013). Meta-analyses since this change have estimated the co-morbidity to be high, with pooled estimates (n=74733) of ADHD prevalence among autism patients of 40% and included studies with estimates of co-occurrence exceeding 70% (Rong et al., 2021). As these new diagnostic standards are propagated, it will be crucial to include co-occurrence in analyses of girls.

### Future Research

Further analysis of income trends in girls with autism and ADHD below 50% FPL and above 400% FPL are necessary. A log(income) analysis was not deemed appropriate in this paper; however, such analyses may be indicated for datasets that include income above 400% FPL.

A U-shaped relationship was detected in ADHD diagnoses in boys near 266% of the federal poverty level. Although outside the scope of this analysis, further investigation of this relationship may yield relevant information for the diagnosis of autism and ADHD.

Finally, the authors recommend that future researchers include the individual races of persons with Hispanic ethnicity. The authors believe that treating Hispanic communities as a racial monolith is detrimental to research, and several of the demographic trends documented in this paper would not be observable without consideration for the full racial diversity of Hispanic girls.

## Conclusion

Girls with autism and ADHD face unique challenges that are affected by SES, race, ethnicity, age, insurance, and family structure. This analysis sought to show that income would have a positive bimodal effect on the diagnosis of autism and ADHD when controlled for the other confounders. This was not the case. Instead, girls have a near-linear relationship between income and ADHD diagnosis, and a slight negative relationship between income and autism diagnosis. Increasing income was associated with slightly lower odds of autism diagnoses. This was heavily influenced by a peak in diagnoses near 138% of the federal poverty limit. The conclusions of this analysis directly contradict preexisting SES and ethnoracial trends in autism and ADHD. In addition, the peak at 138% FPL suggests that Medicaid eligibility may strongly influence income-diagnosis curves for girls and should be further researched. These conclusions are heavily limited by the lack of full income ranges used. Future analyses with datasets that include a wider and more granular range of income are necessary.

## Supporting information

Supplementary STATA File

## Data Availability

All data produced are available online at https://www.census.gov/programs-surveys/nsch/data/datasets.html

